# Understanding the acceptability, barriers and facilitators to implementing the 4CMenB vaccine for the prevention of gonorrhoea in gay, bisexual and other men who have sex with men

**DOI:** 10.64898/2026.06.25.26355673

**Authors:** Roeann Osman, Andrew Jajja, Benjamin Weil, Tom Doyle, Bill Berners-Lee, Fabiana Lorencatto, Hamish Mohammed, Helen Campbell, Shamez N Ladhani, Sema Mandal, Caroline Sabin, John Saunders, Emily Jay Nicholls

**Author notes:** Corresponding author: Roeann Osman, UK Health Security Agency, 61 Colindale Avenue, Colindale, London NW9 5EQ.

## Abstract

**Background:** In November 2023, the Joint Committee on Vaccination and Immunisation advised the UK government to introduce a targeted, opportunistic vaccination programme using 4CMenB in sexual health services to prevent gonorrhoea primarily in gay, bisexual and other men who have sex with men (GBMSM) at higher risk of infection. Evidence on the acceptability of 4CMenB vaccination was required.

**Methods:** Three focus group discussions (FGDs) were conducted with 17 GBMSM aged ≥18 years, resident in England, who selflZlreported bacterial sexually transmitted infection or ≥5 sexual partners in the previous 12-months, alongside one FGD with five sexual healthcare professionals (HCPs). Data were analysed using reflexive thematic analysis, and organised and interpreted using the Vaccine Uptake Continuum and the Social Ecological Model.

**Results:** Acceptability of 4CMenB was high among GBMSM and HCP participants. GBMSM described vaccination as supporting sexual wellbeing and reducing anxiety about gonorrhoea. While the estimated effectiveness (30-35%) was perceived as modest, it did not deter acceptability but reduced willingness to actively seek vaccination. Structural constraints (e.g. limited appointment availability) and restrictive eligibility criteria were identified as barriers to equitable uptake. Community-based delivery models were supported to improve access. HCPs drew on mpox vaccination experience to anticipate implementation challenges and emphasised clear guidance and sustainable resourcing. Additional protection against meningitis was generally a secondary influence.

**Conclusions:** 4CMenB vaccination for gonorrhoea was acceptable to GBMSM and HCPs; however, uptake is likely to depend on ease of access, clear communication, and system-level support. Addressing structural constraints and supporting community-based delivery may help achieve equitable delivery of 4CMenB.

## Background

Gonorrhoea, a sexually transmitted infection (STI) caused by *Neisseria gonorrhoeae*, poses an urgent public health threat due to the emergence of antimicrobial resistance (AMR) (1, 2). In England, gonorrhoea diagnoses have risen over the past decade, increasing from 41,290 reported cases in 2014 to 71,802 in 2024 (3). Gonorrhoea diagnosis rates are disproportionately high among gay, bisexual and other men who have sex with men (GBMSM) (3). Sexual health services (SHSs) across the UK already routinely deliver hepatitis A and B, mpox, and human papillomavirus vaccination for eligible GBMSM alongside other STI, human immunodeficiency virus (HIV) and blood-borne virus (BBV) preventative interventions. In November 2023, the Joint Committee on Vaccination and Immunisation advised the UK government to consider initiating of a targeted, opportunistic immunisation programme in sexual health services using 4-component meningococcal B (4CMenB) vaccine to prevent gonococcal infection primarily in GBMSM at high risk of infection (4). The effectiveness of 4CMenB in reducing the risk of gonorrhoea acquisition has been estimated to be between 33% and 47% in retrospective and prospective observational studies (5). However, evidence from DOXYVAC, a randomised controlled trial assessing the impact of 4CMenB for gonorrhoea prevention, concluded a non-significant relative reduction of 22% (6) and more recently, the GoGoVax randomised controlled trial found no protection against gonorrhoea (7, 8). Nonetheless, modelling studies suggest that even with low or moderate individual-level protection, 4CMenB may confer wider public health benefits by reducing overall gonorrhoea prevalence and antimicrobial use, thereby reducing the burden of AMR (9, 10). 4CMenB is licensed to protect against meningococcal serogroup B (MenB) disease and is expected to provide coverage against most circulating MenB strains causing invasive meningococcal disease, including meningitis and septicaemia, in the UK.

Factors influencing vaccine acceptability and uptake are complex, and likely to be influenced by multiple factors at the individual, interpersonal, organisational and societal levels. Target population and healthcare professional (HCP) support for vaccination are both critical to achieve the intended outcomes of a vaccine programme.

There remains limited understanding of the acceptability of 4CMenB vaccination for gonorrhoea prevention among GBMSM, or the perspectives of HCPs responsible for identifying and vaccinating eligible individuals. We present findings from focus group discussions (FGDs) with HCPs and GBMSM participants to explore perceptions, awareness, accessibility, and acceptability of 4CMenB. We draw on two complementary social science frameworks to inform and structure our analysis (the Vaccine Uptake Continuum) (12) and to interpret findings (the Social Ecological Model (SEM)) (13). The approach allows examination of the interplay of individual and system-level influences on vaccine acceptability and uptake.

## Methods

### Study design

We conducted FGDs with GBMSM and HCPs between Feb/March 2025, using materials co-designed with community-based sexual health organisation The Love Tank.

### Participant identification and recruitment

A call for participation, designed by The Love Tank, was circulated on the social media accounts of Yorkshire MESMAC and The Love Tank. This included a link to a recruitment questionnaire, where potential participants confirmed meeting the inclusion criteria and provided socio-demographic information. We purposively sampled individuals from different age ranges, ethnic groups and geographic regions. HCP participants were identified through professional networks. We obtained consent electronically. GBMSM were eligible if they were cis- or transgender men or assigned male at birth non-binary people who have sex with men, aged ≥18 years, resident in England, with either a self-reported diagnosis of a bacterial STI or ≥5 sexual partners in the last 12 months.

### Setting

FGDs, one each in London, Leeds and online, were conducted with GBMSM. We conducted an online FGD with HCPs employed in SHSs. Remote FGDs were conducted via Microsoft Teams. Each FGD was co-facilitated by two members of the study team (EJN and JS/AJ/RO), or one member of the study team (EJN) and a member from the partnering sexual health community organisation (BW).

### Materials

Members of a Patient and Public Involvement and Engagement panel and The Love Tank reviewed recruitment and participant materials. Topic guides covered similar areas across groups, with relevant sections tailored accordingly, and explored attitudes to gonorrhoea; knowledge and use of STI prevention; perceptions of 4CMenB effectiveness and dosing; and experiences of SHS (see supplementary material A).

### Analysis

RO analysed GBMSM and HCP transcripts separately using reflexive, inductive thematic analysis in NVivo 15 (14). A coding framework was iteratively refined with EJN, JS, FL and HM. Themes were deductively mapped to the Vaccine Uptake Continuum stages: awareness of health threat, availability, accessibility and acceptability (see Supplementary materials B and C). We synthesised findings across GBMSM and HCP discussions by presenting shared concerns and unique considerations from each participant group. SEM was subsequently used to interpret findings.

### Ethics

Ethical approval was granted by the UKHSA Research Ethics and Governance Group (NR0390).

## Results

Seventeen GBMSM (Table 1) and five HCPs (four consultants and one specialist nurse; four women and one man) participated in the focus groups.

**Table 1.** Characteristics of GBMSM participants.

| *Table 1 - Characteristics of GBMSM participants* |  |
| --- | --- |
| **Characteristic** | **Number of participants (n=17)** |
| **Age (years)** |  |
| 18-24 | 3 |
| 25-34 | 6 |
| 35-44 | 4 |
| 45-54 | 4 |
| **Born in UK** |  |
| Yes | 8 |
| No | 9 |
| **Ethnicity** |  |
| Asian or Asian British | 3 |
| Black, Black British, Caribbean or African | 2 |
| Mixed or multiple ethnic groups | 2 |
| White | 10 |
| **Region** |  |
| East of England | 1 |
| London | 8 |
| North West | 1 |
| South East | 1 |
| Yorkshire and the Humber | 6 |
| **Education level\*** |  |
| Currently studying | 1 |
| A-levels or equivalent | 3 |
| Degree or equivalent and above | 12 |

\*Missing data.

Below we present our analysis, structured according to the vaccine continuum domains, enabling identification of barriers and facilitators across the vaccine pathway.

### Awareness of health threat

#### Gonorrhoea

GBMSM participants described gonorrhoea infection as a minor inconvenience or annoyance that temporarily impeded sexual activity but did not cause excessive worry or concern.

> “It’s just an annoying little thing that needs to get away. Get tested, get treated, and forget about it is the way I look at it.” **GBMSM, London**

Vaccination was seen as supporting sexual wellbeing and pleasure, rather than as a response to perceived severe illness. Participants also drew on comparisons between HIV and bacterial STIs; whilst HIV prevention was described as widely discussed and normalised within queer communities, bacterial STIs were considered to be overlooked:

> “We always talk about, ‘are you on PrEP [HIV Pre-Exposure Prophylaxis]? Are you tested?’ But they don’t go into details about what other STIs are there. I feel like most of us take PrEP. We’re all trying to stop getting HIV because I think once you get HIV, you get it, but with the other the STIs, it’s okay. So, no one really talks about the other STIs.” **GBMSM, London**

4CMenB vaccination was seen as an opportunity to rebalance this discourse, normalising conversations about STI prevention. In the context of the availability and uptake of existing biomedical prevention (e.g. PrEP), 4CMenB was often viewed as easy to integrate into current strategies used by participants and their communities. Some GBMSM also expressed motivation to support SHSs, who they perceived to be under strain, and contribute to collective health protection, positioning vaccine uptake as a public good.

GBMSM acknowledged that limited knowledge about bacterial STIs and available prevention strategies could pose a barrier to 4CMenB vaccine uptake. Gaps in accessible information were frequently noted, particularly outside of clinical or queer spaces. Moreover, there may be additional barriers to information for heterosexual-identifying men who have sex with men:

> “I’ve been with DL [down low], discreet men [heterosexual-identifying men who have sex with men], and a lot of them are so misinformed about STIs, and a lot of them don’t go and have PrEP, etc…. They are not going to STI clinics.” **GBMSM, London**

#### MenB meningitis protection

Protection for meningococcal meningitis and septicaemia were seen as secondary benefits rather than key motivators and were, therefore, not the primary “threats” the vaccine provided protection for; however, for some GBMSM it reinforced vaccine acceptability. HCP participants perceived protection against MenB meningitis as a less influential factor in patient decision-making. Although some acknowledged that the dual benefit of protection against meningitis might reassure patients, others suggested it would not be a major determinant of vaccine uptake.

> “it’s a nice added bonus, isn’t it? But I don’t think it’s going to, I’m not sure it would really sell it to a candidate. You could say, ‘and you get extra protection against meningitis, but I think, they’re probably already going to go for it anyway.” **HCP**

### Ensuring accessibility of the vaccine

#### Structural factors

Both participant groups emphasised that structural accessibility would be the largest barrier for patient uptake. Access to face-to-face SHS appointments was frequently cited as limited, which may reduce opportunities for vaccine delivery. This is particularly important given the twolZldose regimen for 4CMenB, as any barriers to attending the first appointment are likely to be compounded when returning for the second dose. In the context of relatively low concern about gonorrhoea acquisition (described in the previous section), the effort required to secure an appointment and navigate overstretched services was often perceived as disproportionate to the anticipated benefit of vaccination. As one GBMSM participant explained, willingness to be vaccinated was contingent on ease of access:

> “…if you’re telling me that I need to make a lot of phone calls, a lot of waiting time, then to wait maybe three months, then I’m like, ‘what is the point?’ **GBMSM, London**

This was particularly so in relation to attitudes around the relatively low estimated effectiveness of the vaccine (described later). A small number of participants noted the possibility to privately pay for access to the vaccine at community pharmacies under its license for Meningitis B, recognising that this could further exacerbate inequalities:

> “I know quite a few people who have gone down that private route to the vaccine […] not everybody has that privilege to be able to pay for the vaccine privately, so just access is really very key here.” **GBMSM, London**

Overall, integrating 4CMenB provision into existing SHSs was regarded as feasible by HCP participants. However, similar to GBMSM participants, HCPs identified multiple structural factors that could facilitate or hinder equitable access to the 4CMenB vaccine. For instance, HCP participants drew on experiences with the rollout of the mpox vaccination, reflecting on how limited appointment availability or regional disparities in service delivery could have an impact on accessibility.

GBMSM participants similarly commented on potential barriers to access, including travel and opening times:

> “It’s hard to get to clinics, and again, when you’re thinking about rural areas, your more socioeconomic, depressed areas, people can’t afford to take time off work to go to the clinic. Clinics generally only open during working hours, so people can’t afford to start with the working hours. People might not be able to afford to get a bus out to the clinic to get it, especially, again, if it’s quite a distance away” **GBMSM, Leeds**

Participants discussed the potential for eligibility criteria to be a barrier to equitable access. While the proposed offer to GBMSM attending SHSs was understood as important for targeting populations at higher risk, several recognised the tacit exclusion of certain groups who may benefit from vaccination. Participants suggested that identity-based eligibility could limit uptake among individuals whose sexual behaviour places them at risk, but who do not align with current eligibility criteria:

> “I think one of the issues that there could be with just marketing the vaccine towards men who have sex with men is there are a lot of men who have sex with men who don’t see themselves as gay or bisexual or what have you, that they are, and then won’t get the vaccine because clinics are really bad at digging information out of people” **GBMSM, Leeds**

Participants also raised concerns that several groups at elevated risk of gonorrhoea were not explicitly named in proposed eligibility criteria, with HCPs noting that their omission could perpetuate inequities in access. As one HCP explained:

> “Prisons, that would obviously have been on my list. Then sex workers would be my other. Particularly a bit like we saw in PrEP, we saw a very big challenge with getting women engaged in PrEP, any female dominated cohorts”. **HCP**

Both GBMSM and HCPs emphasised that achieving equitable access would depend not only on patient willingness but on whether clinicians consistently identified and offered vaccination to those at higher behavioural risk. Participants described how, in practice, eligibility decisions often hinge on individual clinician judgement, leading to inconsistencies between and within services. Although national immunisation guidance states that 4CMenB should be offered to anyone attending SHS with a comparable level of gonorrhoea risk as eligible GBMSM irrespective of gender or sexual identity (15), participants worried that this risklZlbased approach may not be uniformly applied. HCPs therefore stressed the need for clearer national guidance and standardised criteria to support consistent implementation and avoid local or regional disparities in vaccination offer.

> “I don’t think my concern is so much about the eligible patients and the chaos around that, I think it’s the ineligible patients and the chaos and comms around those who are familiar with gonorrhoea that might not fall into the gay/bisexual MSM category.” **HCP**

### Facilitating accessibility through outreach

To address barriers associated with limited SHS capacity and difficulties accessing face-to-face appointments, both participant groups proposed outreach and community-based delivery models as strategies to improve access to 4CMenB vaccination. These approaches were viewed as reducing the effort required to obtain vaccination by bringing services closer to where people already socialise. GBMSM participants emphasised the importance of decoupling vaccination from SHSs and expanding delivery through community organisations. This was framed as a way to normalise vaccination, reduce reliance on overstretched services, and increase convenience:

> “I would really like to see a shift where it’s not just the STI clinic where you get your vaccines. I think community services [should be] being able to give the vaccines. You don’t need to be a nurse to do a vaccine. We found out in COVID [that] anybody can do a vaccine as long as you get the right training.” **GBMSM, Leeds**

HCP participants similarly described outreach delivery as a pragmatic response to access constraints, particularly for populations at higher risk who may not routinely attend SHSs. Some suggested delivering vaccination directly within sex-on-premises venues or other community settings to minimise barriers related to appointment availability:

> “Could you take that to a sex on premises venue, assuming you’ve got all the infection control precautions in place? Would that be a more pragmatic way of doing it? Because if you’re in a sex on premises venue as a gay man, you’re probably more likely to get gonorrhoea than many other conditions, aren’t you? So, that would take it straight to the people that would need it”. **HCP**

HCPs also highlighted the need for sustainable resourcing and workforce planning to ensure that outreach provision could be maintained alongside existing SHS activity, rather than displacing already limited clinic capacity:

> “So, that would take it straight to the people that would need it, but I guess you’d need the commissioning and the funding structures to support that, which they probably wouldn’t be, where there isn’t an excess of money floating around the system.” **HCP**

GBMSM participants highlighted the importance of community-led messaging, including promotion via geospatial dating and social networking applications, to reach populations disengaged from SHSs and increase vaccine access and uptake. HCP participants emphasised the importance of communication and training to support both patients and HCPs, as these were seen as fundamental to ensuring clarity about patient eligibility, and vaccine safety.

### Facilitating consistent and efficient service delivery

HCPs emphasised that ensuring equitable access to 4CMenB would require clear guidance on prioritisation and delivery within SHSs, and highlighted that in the absence of national guidance, decisions about which vaccines to prioritise were often made pragmatically, influenced by time pressures, staffing, and risk urgency. Participants expressed uncertainty about where 4CMenB vaccination should sit within existing vaccine administration hierarchies and whether other vaccines could be co-administered safely. Participants therefore stressed the importance of standardised guidance to support consistent decision-making across clinicians and services:

> “Sometimes you end up prioritising, obviously, the treatment for the STI and then seeing how they feel about all the multiple vaccines and thinking which one to prioritise first. Is it Hep B? Then you come back another day for the mpox, or you have mpox and Gardasil on the other day. I don’t know about the interaction between the vaccinations either, so that’s my knowledge gap. I don’t know what happens if you have Gardasil and [4CMenB] on the same day, or is there a site-specific element?” **HCP**

Perceptions of the opportunistic offer of 4CMenB vaccination at the time of gonorrhoea diagnosis and treatment were mixed among GBMSM participants. Most participants viewed this approach as efficient and pragmatic due to minimising the need for additional appointments, particularly in the context of limited SHS capacity. However, some participants expressed discomfort with linking vaccination to infection treatment, citing negative emotional associations with a gonorrhoea diagnosis or the invasive nature of treatment as potentially undermining acceptability for some individuals:

> “My experience has not been good […] especially on treatment, so if it’s any connection to that, I would say a red flag goes up in my mind.” **GBMSM, Online**

HCP participants drew on experience from mpox vaccination programmes to inform expectations around the implementation of 4CMenB. Early rollout was anticipated to be resource-intensive, requiring additional staff training, and patient communication strategies, which could collectively impact service capacity and appointment availability. However, participants emphasised that once delivery processes become embedded into routine care, vaccination could be delivered more efficiently, and reduce operational burden and improve access in the long term:

> “I think the first couple of months, 2-3 months, are going to be finding feet and managing expectations, and then it will settle into something. It’s just what that something is. I think it’s going to take longer at the beginning, obviously, but once we are, our experience locally has been that the clinic seems to get to grips with things quite quickly and it then gets embedded into your service, and it’s okay”. **HCP**

### Availability of the vaccine

Ensuring a consistent supply chain was a key logistical consideration by HCP participants, alongside a recognition that inconsistent vaccine supplies could have an impact on service provision. Resource limitations, particularly time, cost, and staff shortages were also frequently cited barriers by HCPs:

> “… supply is quite important. There needs to be a steady and consistent supply of vaccine that’s able to get to different parts of the country wherever it’s needed, because disruptions to that cause a bit of chaos in clinics as well.” **HCP**

Several HCPs drew on recent experiences of delivering mpox vaccination to illustrate how limited nursing capacity had directly disrupted vaccine provision:

> “we’ve had to suspend our vaccine clinic because we don’t have enough nursing staff to cover them.” **HCP**

Prioritisation strategies, such as aligning 4CMenB vaccine delivery with other patient appointments (e.g. PrEP) were discussed as potential opportunities to improve SHS efficiency and reduce patient burden. Pilot testing of 4CMenB delivery was viewed as an important step toward maximising successful national implementation, by allowing SHSs to refine delivery processes:

> “a short pilot or plans to test it out in real-world life and see the kind of patient feedback that is gained from trying to implement it.” **HCP**

### Encouraging acceptability of the vaccine

GBMSM and HCP participants described 4CMenB as highly acceptable:

> “I think patients are really going to want it. They all know what gonorrhoea is. A lot of them have had gonorrhoea before. It’s much more tangible for them than HPV [Human papillomavirus] and Hepatitis B, so I think they are going to want it”. **HCP**

One of the primary drivers of acceptability among GBMSM was the very high level of trust in public health messages, scientific expertise and sexual health clinicians:

> “I’ve still got a lot of trust in public health figures, so I think for me, I think probably for a lot of other people who don’t really go deep too much and just do what they can” **GBMSM, London**

However, a small number of GBMSM participants described experiences where their trust had wavered due to inconsistent and conflicting information provided by sexual health clinicians, such as receiving duplicated vaccine offers in previously vaccinated patients. Participants raised concerns about the reliability of information provided and the accuracy of clinical records, highlighting the importance of consistent messaging and accurate documentation to maintain confidence in vaccine delivery. Participants emphasised that while these experiences did not necessarily deter them from vaccination, they introduced uncertainty and frustration, which could erode trust if encountered repeatedly.

HCP participants reported that a previous gonorrhoea diagnosis could facilitate vaccine uptake among eligible service-users by making the risk more tangible and relevant. The vaccine effectiveness estimate of ∼30-35% elicited varied responses. Some GBMSM participants viewed the protection estimates as being disappointing, but not a deterrent to uptake:

> “It’s lower than what I imagined. So, that isn’t like the, ‘oh, whoopee, that’s the silver bullet. No, it’s not. It might hit sometimes.’ It might miss the two other times. I suppose it’s better than nothing, isn’t it?”. **GBMSM, Online**

While vaccine effectiveness appeared to have little impact on acceptability, participants indicated that it may affect the degree of effort they were prepared to undertake to acquire the vaccine. GBMSM participants therefore described themselves as being highly likely to accept the vaccine if offered opportunistically, but to be unlikely to seek it out, particularly given the difficulties accessing SHS appointments described earlier:

> “I do think the efficacy rate would make me not jump and like go out of my way. I definitely would still get it, but lots of other factors would play into how quickly I would go and get it and how much out of my way I would go, because the waiting times for some of the clinics in London are just fucking stupid.” **GBMSM, London**

Participants highlighted the importance of clear and transparent communication about vaccine effectiveness, including setting expectations of partial protection and whether vaccination reduces transmission or disease severity. Limited understanding of partial protection was seen as a risk for disappointment if gonorrhoea occurred after vaccination, potentially undermining trust in 4CMenB and SHSs, particularly if expectations were not clearly communicated at the point of offer. HCPs similarly anticipated that such experiences could reduce willingness to return for booster doses or engage with future vaccination programmes.

GBMSM and HCPs wanted clear information on how quickly protection lasts before waning, and the expected timing of booster doses. Uncertainty around duration of protection was viewed as a barrier, with HCP participants concerned that infections occurring soon after vaccination could create confusion or diminish confidence in the programme.

Several GBMSM participants also emphasised the importance of understanding the evidence underpinning effectiveness estimates, particularly given the modest level of protection:

> “I want to know the efficacy for each dose, and also when does peak protection happen? How many weeks does that take? How long does protection last as well? And also what the vaccine actually does, does it prevent transmission or is it actually just designed to reduce symptoms?” **GBMSM, London**

Transparent communication about safety and potential side effects was also viewed as essential to support informed decision-making and build confidence in the vaccine:

> “The risks and benefits. Are there any potential side effects? How long has the vaccine been in use?” **GBMSM, Online**

GBMSM participants were also asked whether protection against meningitis would impact their decision-making when offered for gonorrhoea prevention. Attitudes toward this were mixed; for some, existing familiarity with the 4CMenB vaccine reassured participants of its safety, whilst others expressed concern about linking a routine childhood vaccine to STI prevention:

> “It’s so weird to hear the two things in the same sentence. I think it would cause a lot of furrowed brows as well. It’s like, what? The thing you get for meningitis, you can get for gonorrhoea?” **GBMSM, Online**

Participants with family members affected by meningitis described this association as a strong motivator for uptake. In contrast, others reported that meningitis was not something they had previously considered relevant to them.

### Contextualisation of results using the SEM

Using SEM, we interpret our findings across multiple interacting levels to illustrate how individual, interpersonal, organisational, and wider community and policy contexts collectively shape vaccine acceptability and uptake (Figure 1). At the individual level, trust in public health institutions influenced willingness to vaccinate among GBMSM participants, as well as feelings of civic duty towards others. Although some participants expressed disappointment about the effectiveness estimate, this did not deter interest in vaccination uptake. However, several questioned how such effectiveness estimate data might be received within their wider communities.

**Figure 1:**
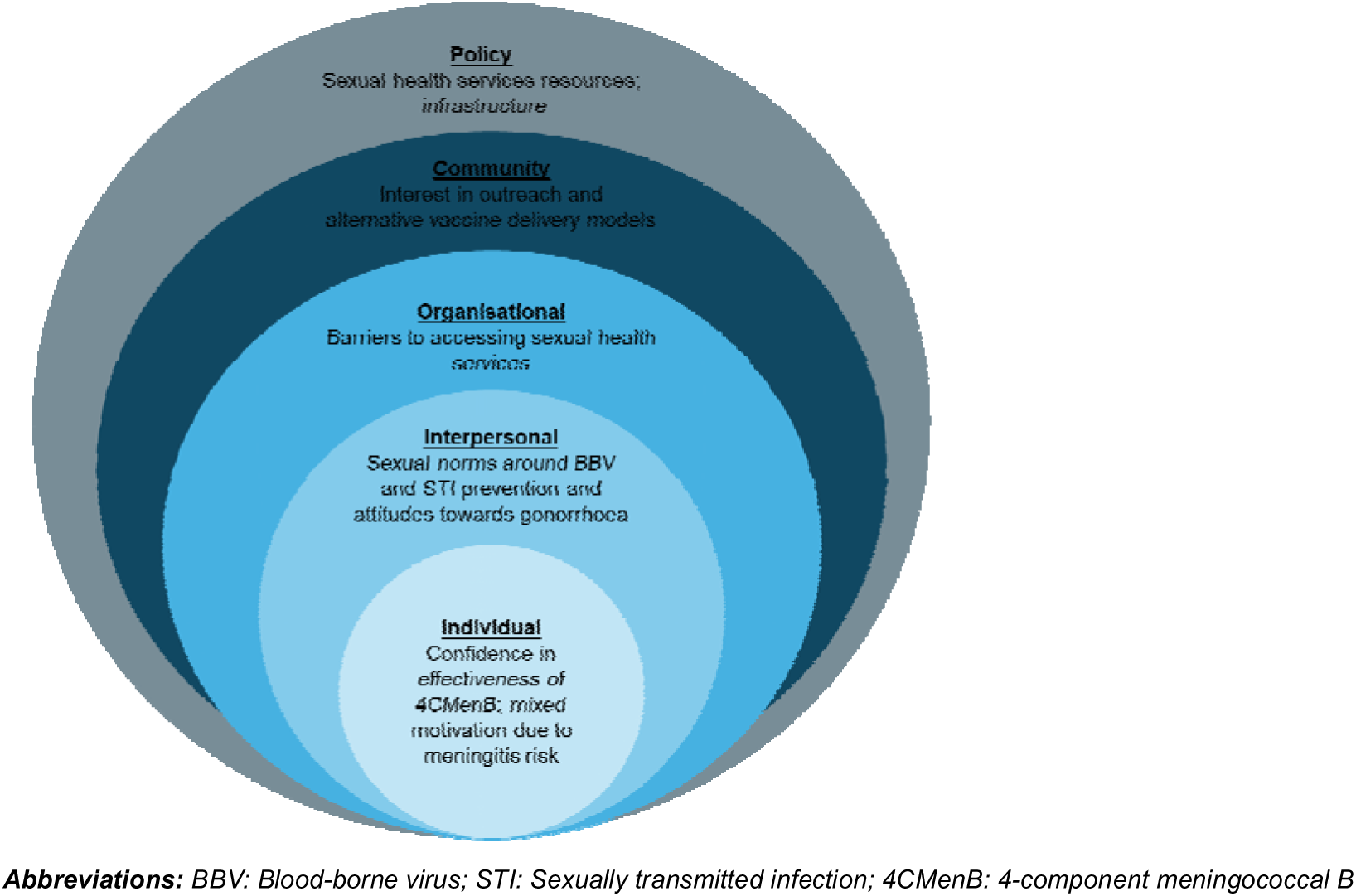
Summary of themes in the Social Ecological Model

At the interpersonal level, social networks and peer norms shaped attitudes towards STI and BBV prevention. While HIV prevention was widely discussed, bacterial STIs were felt to receive less focus. At the organisational level, HCPs identified structural barriers including reduced SHS capacity, resource constraints, and regional service provision disparities. While integration into SHS delivery was seen as feasible, adequate staffing, funding, and training were viewed as essential. At the community and policy levels, outreach initiatives and inclusive eligibility criteria were identified as critical to ensuring equitable access. The success of mpox vaccination campaigns demonstrated the value of community partnerships and flexible delivery models, suggesting similar strategies could support 4CMenB implementation.

## Discussion

This study explored attitudes towards 4CMenB vaccination for gonorrhoea prevention among GBMSM and HCPs. Overall, acceptability of 4CMenB was high, with participants expressing strong willingness to receive or deliver the vaccine, driven by its potential to enhance sexual wellbeing and reduce gonorrhoea diagnoses. Participants characterised 4CMenB as a straightforward prevention strategy with potential synergy with other STI and BBV interventions. Participants emphasised the importance of clear communication to promote 4CMenB and encourage uptake. However, participants consistently emphasised that uptake would be contingent on ease of access. Despite enthusiasm for vaccination, limited SHS capacity, inconsistent regional provision, and restrictive eligibility criteria were perceived as barriers to equitable delivery and uptake. Community-based and outreach delivery models were proposed as possible solutions.

Our findings align with existing evidence that frames vaccine acceptability as shaped by intersecting behavioural, social, and structural determinants rather than individual motivation alone (17). Previous qualitative research exploring vaccine attitudes among GBMSM, such as studies of human papillomavirus, mpox, and COVID-19 vaccination, highlighted the role of trust in public health messaging and science (18), perceived personal relevance (19, 20), and improving patient access through using outreach services to facilitate vaccine uptake (21, 22).

Similar themes have also been identified in qualitative research on doxycycline post-exposure prophylaxis (doxyPEP), where acceptability was shaped by perceived relevance to sexual risk, supporting individual and collective health outcomes and confidence in HCP advice (23). Consistent with evidence from COVID-19 vaccination rollouts, both GBMSM and HCPs emphasised the importance of flexible and community-based delivery models (24). DoxyPEP qualitative research similarly highlighted the crucial role of HCPs in tailoring and communicating prevention messages, alleviating patient uncertainty, and framing prevention in ways that resonate with patient experiences, reinforcing the importance of clinical trust and consistent messaging (25).

It is important to note that participant views were formed in the context of the observational evidence available at the time, which suggested that 4CMenB might offer modest protection against gonorrhoea, and prior to a subsequent high-profile meningitis outbreak in the UK (26), which may since have altered perceptions of the value of meningococcal protection. Following the announcement of the GoGoVax trial results showing no efficacy of 4CMenB on gonorrhoea incidence (8), participant acceptability may now be lower. Nonetheless, the emphasis participants placed on trust, communication, and the need for comprehensive prevention options remains highly relevant.

### Implication for policy and practice

Our findings have important implications for the delivery of 4CMenB as well as other sexual health vaccines. Effective communication strategies should prioritise clarity regarding eligibility, dosing, side-effects, and duration of protection while addressing potential misconceptions about lifelong immunity. Ensuring that communication and outreach strategies are inclusive and diverse will be essential to support equitable engagement and uptake alongside structural efforts to improve service access. Equitable access will be essential to avoid amplifying existing health inequalities (27). Embedding outreach and community-based delivery, such as vaccination at queer venues or events, can prioritise inclusivity and avoid replicating inequalities observed in sexual health care.

Consistent delivery will depend on adequate service availability, clear workflows and clinical guidance to support risk-based eligibility and prioritisation. Standardised staff training and communication tools will be needed to ensure uniform delivery within and across sites, especially given previous lessons from mpox and PrEP delivery.

### Future research

Future research should explore acceptability of 4CMenB vaccination among other populations, such as sex-workers. There is also a need to examine how structural inequalities, such as socioeconomic deprivation and ethnicity, shape vaccine access and uptake. Quantitative evaluation of early implementation phases could help identify inequities in vaccine offer and uptake, informing more inclusive delivery strategies, while also providing early real-world evidence on vaccine effectiveness.

### Strengths and limitations

Our study contributes novel insights into the acceptability of the 4CMenB vaccine for gonorrhoea prevention. We included participants across a range of ages, ethnicities, and geographic regions, thus enhancing the transferability of findings. However, our participants were predominantly White, which limits insight into how communities of colour, some of whom may have greater medical mistrust and express heightened concerns about new interventions (16), may perceive and engage with 4CMenB vaccination.

A few GBMSM participants had professional experience in sexual health or community outreach and participated in a personal capacity. Their contributions occasionally drew on this expertise, particularly regarding service delivery and workforce considerations, offering useful implementation insight while not necessarily reflecting the views of all GBMSM service-users. Participants were generally highly educated and health literate, which limits the representativeness of perspectives from more marginalised groups or those less engaged with SHSs. This may have influenced findings related to trust in services and vaccination and understanding of vaccine effectiveness. As we recruited through community-based sexual health organisations, our participants were also more likely to have access to, and familiarity with, the issues discussed in the FGDs.

As the research was conducted prior to national programme rollout, reported attitudes reflect anticipated rather than actual experiences or knowledge of vaccine access and delivery and should be interpreted in the context of the observational evidence available at the time.

## Conclusions

Our study demonstrates high theoretical acceptability of 4CMenB vaccination among GBMSM and HCPs, with enthusiasm tempered by structural barriers and uncertainties regarding equitable access. Addressing multilevel determinants, through tailored communication, community partnership, and resourced service delivery, will be essential to achieving equitable and sustained vaccine uptake.

## Data Availability

Anonymised data may be made available from the corresponding author upon reasonable request and subject to ethical approval.

## Acknowledgements

We would like to thank The Love Tank and Yorkshire MESMAC for supporting recruitment for this study. The authors wish to thank all the participants who took part in this study. We acknowledge members of the National Institute for Health and Care Research Health Protection Research Unit (NIHR HPRU) in Blood Borne and Sexually Transmitted Infections (BBSTI) Steering Committee: Professor Caroline Sabin (HPRU Director), Dr John Saunders (UKHSA Lead), Professor Catherine Mercer, Professor Gwenda Hughes, Dr Hamish Mohammed, Professor Greta Rait, Dr Ruth Simmons, Professor William Rosenberg, Dr Tamyo Mbisa, Professor Rosalind Raine, Dr Sema Mandal, Dr Rosamund Yu, Dr Samreen Ijaz, Dr Fabiana Lorencatto, Dr Rachel Hunter, Dr Kirsty Foster and Dr Mamooma Tahir.

## Author contributions

JS, CS: Funding acquisition.

JS, SM, HC, SNL: Conceptualisation.

JS, AJ, EJN, FL: Methodology.

RO: Formal analysis, Writing – original draft.

EJN, AJ, BW, JS, RO: Investigation.

EJN: Supervision.

BBL, AJ: Project administration.

All authors: Writing – review and editing.

## List of abbreviations

4CMenB: 4-component meningococcal B
AMR: Antimicrobial resistance
BBV: Blood-borne virus
DoxyPEP: Doxycycline post-exposure prophylaxis
FGD: Focus Group Discussion
GBMSM: Gay, bisexual and other men who have sex with men
HCP: Healthcare professional
HIV: Human immunodeficiency virus
MenB: Meningococcal type B
PrEP: Pre-exposure prophylaxis
SEM: Social Ecological Model
SHS: Sexual health service
STI: Sexually transmitted infection

