## Supplementary materials for "Understanding the acceptability, barriers and facilitators to implementing the 4CMenB vaccine for the prevention of gonorrhoea in gay, bisexual and other men who have sex with men"

### Supplementary material A: Topic guides

GBMSM focus group topic guide

**Introduction**

- Welcome and introduction of the facilitator
- Explanation of the purpose of the focus group
- Explanation of confidentiality and consent (including that this cannot be guaranteed due to nature of group discussion)
- Ground rules for discussion (e.g., respect for opinions, one person speaks at a time, do not share anything said in the group)
- Ice breaker: participants preferred (pseudo)name and reason why they decided to join the group

**Gonorrhoea**

- Does gonorrhoea impact the GBMSM community
  - Is it something people worry or are concerned about (e.g. catching/spreading it)?
  - Is it a big issue within this community?
  - What measures do people take to protect themselves?

**Introducing the vaccine**

As you know, for this study we’re interested in knowing your views on a gonorrhoea vaccine and how gay, bisexual and other men who have sex with men might feel about it, what might influence decisions about taking it and how we might communicate about it to this community. So in the next section we’re going to ask you some questions about vaccines in general and then about this vaccine in particular.

This vaccine we are discussing primarily protects against the bacteria that causes meningitis and sepsis, and it is already used in the UK as part of the childhood vaccination programme. There is evidence the vaccine can also provide some (but not full) protection against gonorrhoea. The vaccine is recommended to be given through sexual health services to GBMSM at increased risk of gonorrhoea infection. It may require two doses, 6 months apart. You might receive a vaccine on diagnosis or during a clinic appointment. We are still waiting for a government policy decision on programme and details of the rollout of the programme would need to be determined.

**Vaccination**

- Have you accessed vaccines through sexual health before (e.g. HPV, Mpox, HAV, HBV)?
  - What were your experiences?
- What do you think about the idea of a vaccine for gonorrhoea?
  - What about other gay, bisexual or other men who have sex with men?
  - In relation to other ways of reducing risk?
- What would be important to know about the vaccine to when deciding to get vaccinated?
  - Allow participants to put forward answers and explore each area generally before providing specific information to discuss

| Effectiveness | - The vaccine does not provide complete protection (some people may still get infected) - The vaccine gives around 35% protection against gonorrhoea (1 in 3 people vaccinated might still develop infection) - Is there a minimum level that is acceptable? |
| --- | --- |
| Safety | - The vaccine is licensed for adults and has been given to millions of individuals safely without any serious health events. - The vaccine is **not** licensed for the use of protection against gonorrhoea. |
| Delivery | - There would be two doses. - Having the vaccine on diagnosis of gonorrhoea might mean receiving treatment for gonorrhoea (an injection) and the vaccine at the same time. |
| Duration | - Level of protection offered by the vaccine decreases over time. - The vaccine gives protection for around 3 years. - There is a potential offer of a booster dose after 3 years. - Is there a minimum time of protection that is acceptable? |

- As we mentioned, this vaccination was originally designed for protection against meningococcal bacteria, which causes meningitis and sepsis, so would provide high protection for these.
  - Would that influence your decision around vaccination?
  - Is meningococcal disease something the GBMSM community is aware of or concerned about?
  - There have been outbreaks in this community, would that influence your decision-making about this?
- How do you think GBMSM, and the wider community will respond to the offer of this new vaccine programme?
  - Do you think people will want it?
  - What might people’s motivations be?
    - Individual protection
    - Community protection (even if individual coverage is 35%)?
- Could other people influence decisions about vaccination?
  - Would friends or partners having been vaccinated influence your decision?
  - What role might community leaders or influencers play?
  - Are there any other social or cultural influences in your communities (e.g. religion)?

**Implementation**

- If the vaccine were to be rolled out and offered to the GBMSM, in your opinion, what could be done to help support its uptake in the community?
- How would you like to hear about the vaccine?
  - What would be the best way of advertising to the community?
  - Who do you want to hear it from, and how would you like to receive this information?
  - What kind of information would be useful?
  - How would you like a healthcare professional to discuss this with you?
  - How confident do you think people are in understanding information about vaccines and their purposes?
- What would make the GBMSM community feel heard and valued in this process?

**Closing**

- Anything we haven’t spoken about that you’d like to add
- Summary of key points discussed
- Thank participants for their time and valuable insights
- Information on next steps and how the feedback will be used

HCP focus group topic guide

**Background**

The UK Joint Committee on Vaccination and Immunisation (JCVI) have recommended a new vaccination programme primarily targeted at gay, bisexual and other men who have sex with men (GBMSM) who are at increased risk of gonorrhoea infection. The programme uses the existing 4CMenB vaccine which is currently being delivered routinely in the UK for immunisation against meningococcal group B disease. Although the vaccine primarily protects against meningococcal disease, there is evidence it can also provide protection against gonorrhoea.

**Introduction**

- Welcome and introduction of the facilitator
- Explanation of the purpose of the focus group
- Brief overview of background information
- Explanation of confidentiality and consent (including that this cannot be guaranteed due to nature of group discussion)
- Ground rules for discussion (e.g., respect for opinions, one person speaks at a time, do not share anything said in the group)
- Ice breaker: participants preferred (pseudo)name and reason why they decided to join the group

**Vaccination**

- What is your understanding of the vaccine and its use for the protection of gonorrhoea?
  - Are you aware of current thinking and research behind the vaccine
    - Would this influence how you felt about delivering the vaccine
  - To what extent does this make sense as an intervention
  - Awareness of eligibility criteria and whether it makes sense
- How would this vaccination programme fit with current practice?
  - To what extent do you see this as their responsibility professionally
  - To you think delivery in SRHS is the right place for it
- How do your colleagues view the new vaccine programme and the items we discussed?
  - Is there a general consensus or differing opinions
- How do you think service users and the community will respond to this proposed vaccine programme?
- What do you think the benefits of vaccination would be?
- For clinics
- For service users
- Where would it fall in terms of priority?
  - clinical priority, service priority, etc.

**Implementation**

- What resources are needed for implementing this programme?
  - Staff, time, infrastructure
  - To what extent are these resources in place and what is missing
  - How would this fit into current service provision
- What additional resources, training or support is needed to effectively implement the new vaccine programme?
  - Gaps in skills and/or knowledge
  - Training materials, preferred formats
- How confident would you be implementing this programme?
  - How easy or difficult would it be
  - Would it be easy to identify eligible cohort
- Are there any lessons learned from successful implementation of other similar programmes?
- How should feedback be collected from healthcare professionals during the implementation of this new vaccine programme?

**Closing**

- Summary of key points discussed
- Thank participants for their time and valuable insights
- Information on next steps and how the feedback will be used

| **Domain name** | **Theme name** | **Theme definition** | **Code name** | **Code description** |
| --- | --- | --- | --- | --- |
| **Awareness of health threat** | Attitudes to  BBSTIs | Experiences and feelings about BBSTI acquisition and prevention measures | Experiences and attitudes towards gonorrhoea infection | Experiences and attitudes towards gonorrhoea infection |
|  |  |  | Individual responsibility towards sexual health | Importance of managing and protecting your sexual health |
|  |  |  | Lack of knowledge | Lack of BBSTI knowledge and prevention methods |
|  |  |  | STI preventions used | Other STI prevention methods used (e.g., doxyPEP and vaccines) |
|  |  |  | STIs vs. HIV | Expected risks associated with STI acquisition vs. HIV |
|  | Attitudes to Meningitis | Attitudes towards use of meningitis vaccine | Meningitis protection | Attitudes and emotions associated with vaccination protection against meningitis |
|  | Good citizenship | Positives associated with 4CMenB use at the population level | Caring about the NHS (e.g., cost savings for SHSs and NHS) | The impact of individual 4CMenB uptake on the health system |
|  |  |  | Caring about the community (e.g., feelings of duty towards your community) | The impact of individual 4CMenB uptake on the collective health of GBMSM populations |
| **Ensuring accessibility of the vaccine** | Access and uptake | The barriers and facilitators which contribute to reduced or increased access and uptake | Use of alternative venues and personnel to supplement SHS 4CMenB vaccine delivery | Use of localised and accessible sites and personnel to deliver vaccine administration |
|  |  |  | Access to SHSs | The challenges associated with SHS access (e.g., clinic distance, lack of appointments, clinic open hours, exclusive online access) which contribute to reduced opportunity for face-to-face interaction with clinic staff |
|  |  |  | Communication messages and strategies to increase access, engagement, uptake and knowledge | Approaches to increase knowledge about 4CMenB vaccination to facilitate access and uptake (i.e., what information do people need to know and how do we communicate and disseminate this information to them) |
|  |  |  | 4CMenB vaccine eligibility | Potential for decreased access and uptake among underserved populations (e.g., transgender people, HI-MSM, non SHS-attendees) |
|  |  |  | Administration of treatment and prevention simultaneously | How patients feel about concurrent administration of gonorrhoea treatment and 4CMenB during SHS appointment |
| **Encouraging acceptability of the vaccine** | Trust | Feelings of trust or mistrust | Trust in public health and/ or science | Trust in public health and/ or scientific research advice |
|  |  |  | Trust in healthcare and clinicians | (Mis)trust towards sexual health clinician advice |
|  | Duration of coverage | Thoughts and feelings about the estimated duration of coverage | Forgetting to book/ attend 4CMenB booster appointments | Potential to forget vaccine coverage duration or forget to book 4CMenB booster appointments |
|  |  |  | People assuming it offered protection longer | Impact of patients assuming protection after vaccine has waned |
|  |  |  | Lack of trust due to not knowing what vaccines people have had | Mistrust of sexual health clinicians due to not knowing what vaccines people have had |
|  |  |  | Adequate | Satisfaction with duration of vaccine coverage |
|  | Effectiveness estimate | Emotional responses and understanding of vaccine effectiveness at averting gonorrhoea acquisition | Disappointment | Dissatisfaction with vaccine effectiveness estimate |
|  |  |  | Good enough | Satisfaction with vaccine effectiveness estimate |
|  |  |  | Confidence in effectiveness estimate | Confidence in the reported effectiveness estimate and studies used to generate data |
|  | Vaccine safety | Attitudes on meningitis vaccine safety profile | Safety | Increased confidence in vaccine due to use as part of the routine child immunisation schedule increases confidence |
|  | Vaccine hesitancy | Reluctance or hesitance to vaccination | Impact of vaccine hesitancy (following COVID-19 pandemic) | The effect of increased vaccine scepticism following COVID-19 pandemic on 4CMenB uptake |
|  | Supporting enjoyment | Supporting enjoyment | Supporting enjoyment | Potential of 4CMenB to support fulfilling and enjoyable sex lives/ sexual wellbeing |

### Supplementary material B: Identified themes and associated codes from GBMSM discussions mapped on vaccine continuum domains.

**Abbreviations:** BBSTIs: Blood-borne viruses and sexually transmitted infections; 4CMenB: 4-component meningococcal B; STI: Sexually transmitted infections; HIV: Human immunodeficiency virus; NHS: National Health Service; SHSs: Sexual health services; doxyPEP: Doxycycline post-exposure prophylaxis; GBMSM: Gay, bisexual and other men who have sex with men; HI-MSM: Heterosexual-identifying men who have sex with men

### Supplementary material C: Identified themes and associated codes from HCP discussions mapped on vaccine continuum domains

| **Domain name** | **Theme name** | **Theme definition** | **Code name** | **Code description** |
| --- | --- | --- | --- | --- |
| **Awareness of the health threat** | Patient attitudes and motivations | Perceptions of patient attitudes and motivations | Experiences and attitudes towards gonorrhoea infection | Perception of patient experiences and attitudes towards gonorrhoea infection |
|  |  |  | Meningitis protection | Attitudes and emotions associated with vaccination protection against meningitis |
| **Availability of the vaccine** | Vaccine delivery | Considerations for implementing a new vaccine | Vaccine supply chain | Ensuring a consistent supply of vaccine to services |
| **Ensuring accessibility of the vaccine** | Access | The barriers and facilitators which contribute to reduced or increased access | Access to SHSs | The challenges associated with SHS access (e.g., clinic distance, lack of appointments, use of non-NHS providers) |
|  |  |  | 4CMenB vaccine eligibility | Potential for decreased access and uptake among underserved populations (e.g., transgender people, HI-MSM, non SHS-attendees) |
|  | Vaccine delivery | Considerations for implementing a new vaccine | Use of alternative venues and personnel to supplement SHS 4CMenB vaccine delivery | Use of localised and accessible sites and personnel to deliver vaccine administration |
|  |  |  | Integration into an existing service | How well the vaccination programme can be embedded into existing SHSs |
|  |  |  | Variation in service provision by HCPs | Impact of clinician’s practice on offer and uptake of vaccine |
|  |  |  | Resource shortages (e.g., time, cost and staff) | Impact of resource shortages which could impact success of vaccine programme |
|  |  |  | Communication/ information strategies for patients | Approaches to increase patient knowledge about 4CMenB vaccination to facilitate access and uptake (i.e., what information do people need to know and how do we communicate and disseminate this information to them) |
|  |  |  | Experience of delivering other vaccination programmes | Clinician experience in delivering vaccinations in SHSs |
|  |  |  | Training for HCPs | Approaches to increase clinician knowledge about 4CMenB vaccination to facilitate access and uptake (i.e., what information do people need to know and how do we communicate and disseminate this information to them) |
|  |  |  | Prioritising vaccines | Timing vaccine delivery to improve patient access and reduce vaccine burden |
| **Encouraging acceptability** | Vaccine delivery | Considerations for implementing a new vaccine | Pilot testing | Piloting vaccination programme to support widescale national implementation |
|  | Perception of vaccine estimate | Perceptions about effectiveness estimate | HCP feelings about effectiveness estimate | HCP feelings about effectiveness estimate |
|  | Patient attitudes and motivations | Perceptions of patient attitudes and motivations | Disappointment with effectiveness estimate | Dissatisfaction with vaccine effectiveness estimate |
|  |  |  | Satisfaction with effectiveness estimate | Satisfaction with vaccine effectiveness estimate |
|  |  |  | Impact of patient vaccine hesitancy/ reluctance (following COVID-19 pandemic) | The effect of increased vaccine scepticism following COVID-19 pandemic on 4CMenB uptake |
|  |  |  | People assuming it offered protection longer | Impact of patients assuming protection after vaccine has waned |

**Abbreviations:** 4CMenB: 4-component meningococcal B; HCPs: Healthcare professionals; HI-MSM: Heterosexual-identifying men who have sex with men; NHS: National Health Service; SHS: Sexual health service
